## Supplementary figures for "Statistical examination of shared loci in neuropsychiatric diseases using genome-wide association study summary statistics"

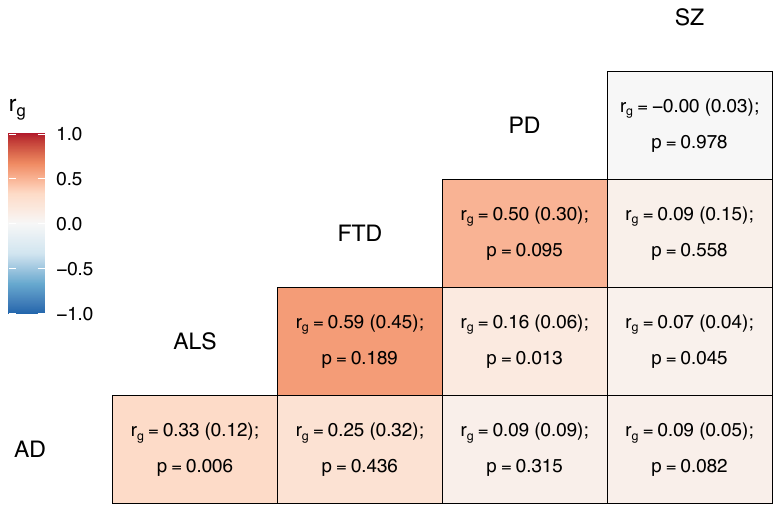

Figure S1. Genome-wide genetic correlation estimates between all trait pairs

The heatmap displays genetic correlations (r_g_) each tile is labelled with the r_g_ estimate and with its standard error in parentheses, alongside the p-value. AD = Alzheimer’s disease, ALS = amyotrophic lateral sclerosis, FTD = frontotemporal dementia, PD = Parkinson’s disease, SZ = schizophrenia.

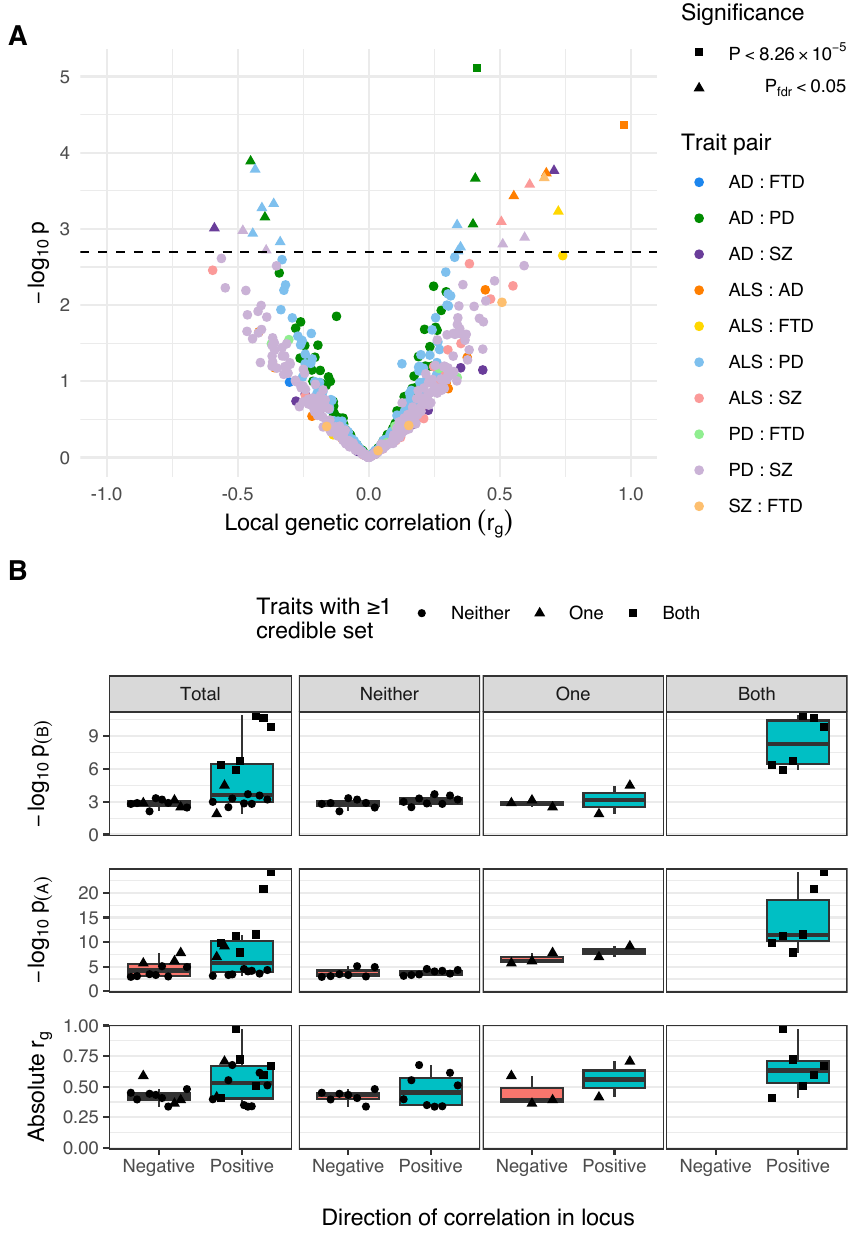

Figure S2. Comparison of positively and negatively correlated genetic loci

**Panel A:** Distribution of genetic correlation estimates (r_g_) and p-values across trait pairs in local genetic correlation analysis. Trait pairs are denoted by colour and comparisons passing defined significance thresholds are denoted by shape (square for a strict Bonferroni threshold and triangle for a false discovery rate (FDR) adjusted threshold); the hatched line indicates the threshold p-value above which P_fdr_ <0.05. **Panel B:** Boxplots comparing FDR-significant loci with positive and negative local genetic correlations. Sub-panels are split row-wise by comparison measure: ‘-log_10_ p_(A)_’ and ‘-log_10_ p_(B)_’ show the p-value for the most significant variant in a given locus for each of the two analysed traits, with ‘A’ being the trait with more-significant and ‘B’ less-significant top variant; ‘Absolute r_g_’ shows the absolute correlation coefficient for the local genetic correlation between trait pairs in a given locus. The leftmost column of sub-panels displays the results across all significant loci and remaining columns split data by the number of traits in the pair with at least one fine-mapping credible set identified at that locus.

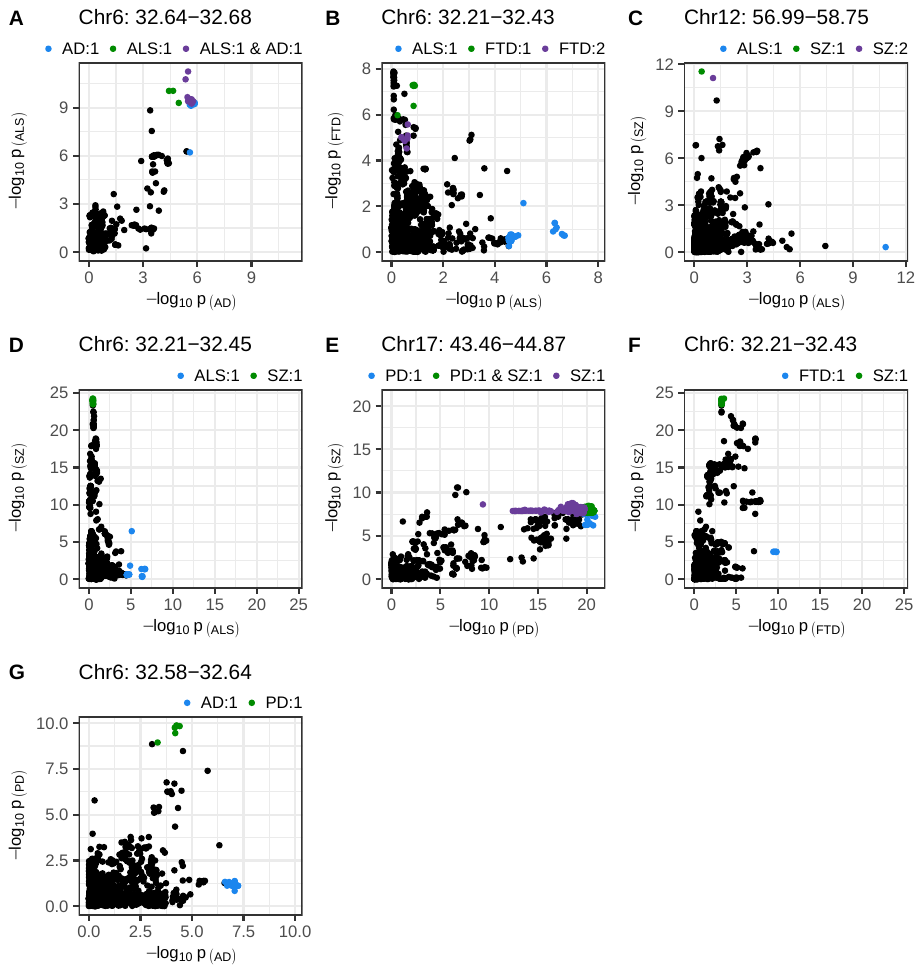
Figure S3. SNP-wise p-value distribution between trait pairs in comparisons where colocalisation analysis suggested a causal variant in both traits

Colocalisation analysis supported the shared variant hypothesis for the comparison in panel A, and the presence of distinct variants for each trait in all other panels (see Table 3). Colouring indicates fine-mapping credible sets assigned to SNPs across the traits compared; the legend above each panel is in the format 'Trait: credible set number'. The genomic position range shown above each panel is in Mb. AD = Alzheimer’s disease, ALS = amyotrophic lateral sclerosis, FTD = frontotemporal dementia, PD = Parkinson’s disease, SZ = schizophrenia.

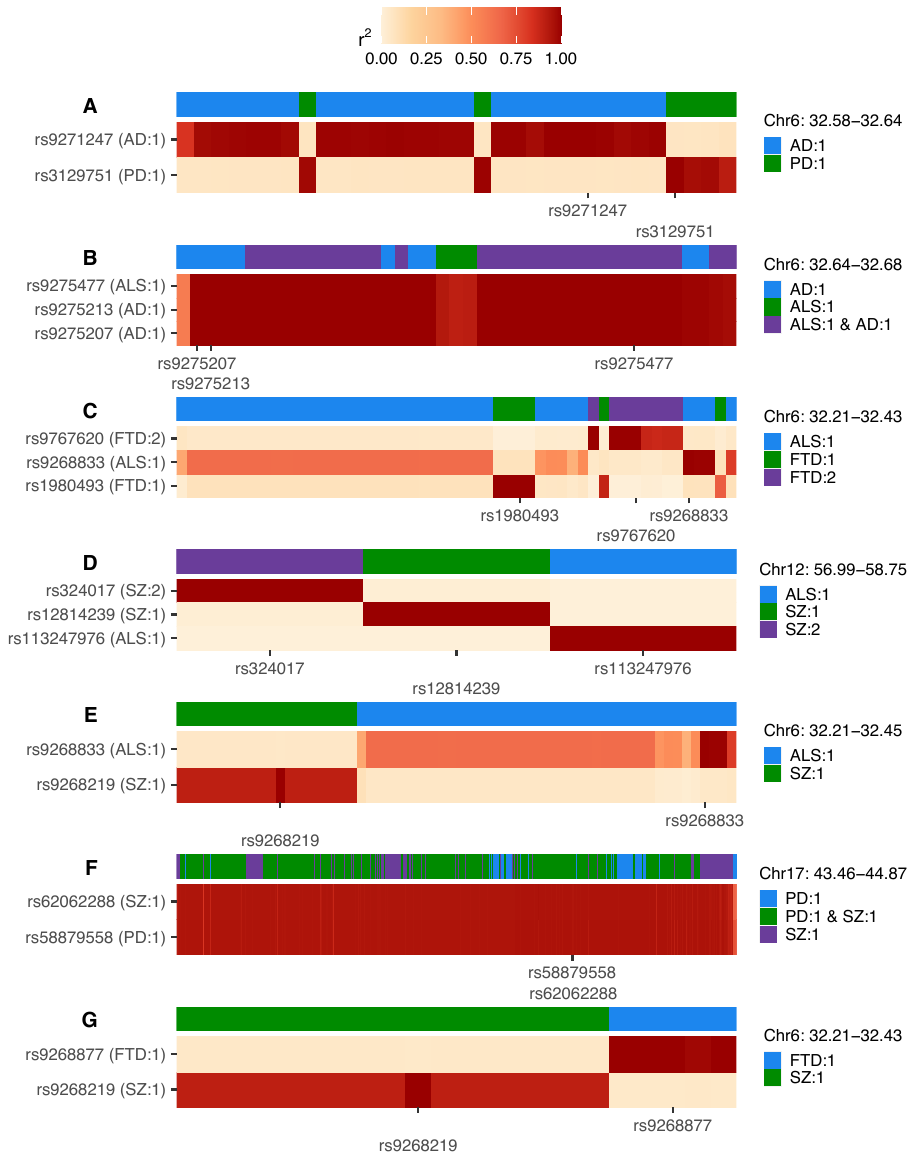
Figure S4. Heatmaps of linkage disequilibrium (LD) in the 1000 Genomes European reference population across variants assigned to any credible set during univariate fine-mapping of trait pairs

LD is shown relative to the SNPs with the highest posterior inclusion probability (PIP) for each credible set, displaying all top PIP SNPs when ties occur. The y-axis splits by top PIP SNPs and the x-axis displays SNPs ordered by genomic position, marking only the positions of the top PIP SNPs. Credible set assignments for each variant are shown in the colour bar at the top of each panel and for the top PIP SNPs in the y-axis label; these are annotated in the format: ‘trait: credible set number’. The genomic range indicated at the top right of each panel refers to the positions spanned across all SNPs analysed and is in Mb. AD = Alzheimer’s disease, ALS = amyotrophic lateral sclerosis, FTD = frontotemporal dementia, PD = Parkinson’s disease, SZ = schizophrenia.

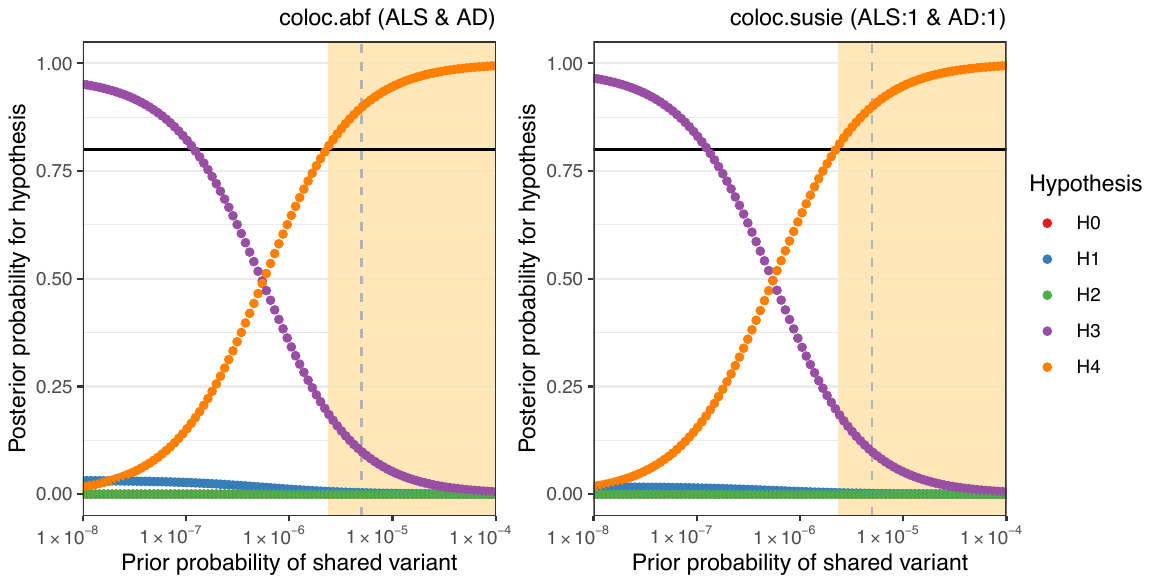

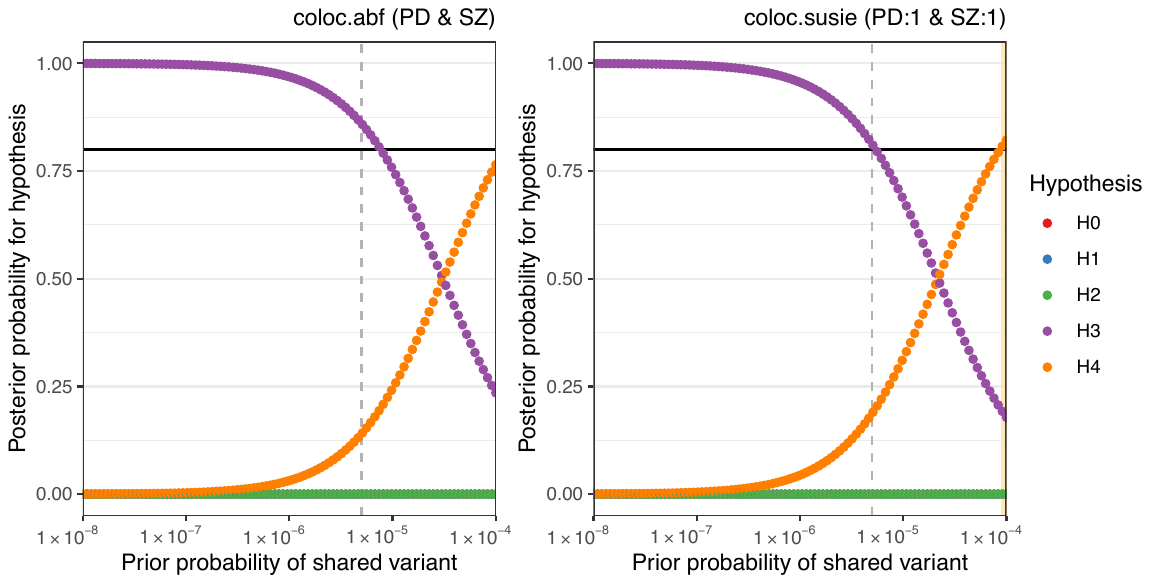
Figure S5. Sensitivity of colocalisation analysis to the prior probability of a shared variant between traits

The upper panels display analysis at Chr6:32629240-32682213 between amyotrophic lateral sclerosis (ALS) and Alzheimer’s disease (AD). The lower panels are for Chr17:43460501-44865832 between Parkinson’s disease (PD) and schizophrenia (SZ). Panels labelled ‘coloc.abf’ display analysis across all SNPs in the region and ‘coloc.susie’ indicates analysis across the SNPs within the pair of fine-mapping credible sets identified across trait pairs. Plot points indicate posterior probability of each hypothesis (H0 = no causal variant for either trait, H1 = variant causal for the trait one, H2 = variant causal for trait two, H3 = distinct causal variants for each trait, H4 = a shared causal variant between traits), according to the prior probability of H4. The vertical hatched line indicates the prior H4 probability defined for the reported analysis; the black horizontal line indicates the defined threshold for acceptance of H4: posterior H4 probability > 0.8. Cream shading of the plot area indicates prior H4 probabilities which yield a posterior probability of H4 above the threshold.
